# COVID-19 and mortality risk in patients with psychiatric disorders

**DOI:** 10.1101/2021.04.08.21255046

**Authors:** George Kirov, Emily Baker

## Abstract

COVID-19 has already caused the deaths of over 2.5 million people worldwide. Patients with certain medical conditions and severe psychiatric disorders are at increased risk of dying from it. However, such people have a reduced life expectancy anyway, raising the question whether COVID-19 incurs a specific risk for such patients for dying, over and above the risk of dying from other causes.

We analysed the UK Biobank data of half a million middle-aged participants from the UK. From the start of 2020 up to 24^th^ January 2021, 894 participants had died from COVID-19 and another 4,562 had died from other causes. We demonstrate that the risk of dying from COVID-19 among patients with mental health problems, especially those with dementia, schizophrenia, or bipolar disorder, is increased compared to the risk of dying from other causes. This increase among patients with severe psychiatric disorders cannot be explained solely by the higher rate of diabetes or cardiovascular disorders.

## Introduction

The epidemic of COVID-19 was caused by the Severe acute respiratory syndrome coronavirus-2 (SARS-CoV-2). The infection leads to severe respiratory tract infections with a high mortality, mostly in the elderly. COVID has a higher mortality than similar respiratory virus infections, with a 2.9 times higher in-hospital mortality for COVID-19 compared to influenza (Piroth et al, 2020). Many risk factors for death are similar, but patients with COVID-19 were more frequently obese or overweight and had diabetes and hypertension, than those with influenza.

The COVID epidemic has cost the lives of over 2.5 million people worldwide (as of March, 2021). Already in the very early stages it became clear that the risk was higher among the elderly, among men, those who were obese, or had hypertension or diabetes (e.g. Zhou et al, 2020; Goodman et al, 2020). Later, it was shown that people from more deprived social backgrounds and those belonging to ethnic minorities were also at an increased risk. Pre-existing medical conditions, such as congestive heart failure, dementia, chronic pulmonary disease, liver or renal disease, and cancer have also been shown to increase the risk (e.g., Cho et al, 2021).

We wanted to explore whether patients with psychiatric disorders had an increased risk of dying from COVID-19. Severe psychiatric disorders, such as psychosis and bipolar affective disorder shorten the life expectancy by one to two decades (Lawrence et al, 2013). Such people can also share some of the risk factors identified for COVID-19, such as increased social deprivation and medical comorbidities, thus placing them at potentially higher risk. An increased risk for COVID-19 related death among patients with mental health problems has indeed been identified (Toubasi et al, 2021).

However, it is also well known that life expectancy in general is reduced for men, for older people, among ethnic minorities, people from deprived social background, and those with medical co-morbidities and increased BMI. Some of these risk factors are also highly correlated with each other. Thus, in an analysis from the UK Biobank (UKBB) Elliott et al, (2021) pointed out that effect of some of the reported risk factors, such as BMI, could be accounted for by age, sex and ethnicity. This team also compared the data for people who died of other causes during the same period and noticed that some of the comorbidities were similar for COVID-19 and non-COVID-19 deaths. By that time 459 UKBB participants had lost their lives due to COVID-19.

We wanted to answer the question whether psychiatric disorders are risk factors for dying from COVID-19 over and above the risk for such people dying from other causes.

## Methods

We used data from the UKBB on half a million participants from the UK, which was made available for research on COVID-19. The UKBB is particularly suitable for this research, as the population is now middle to old-age and the researchers have accumulated extensive information on baseline characteristics and medical outcomes. Our data download on COVID-19 was completed on 6^th^ March 2021, after the latest peak of deaths caused by the pandemic. From the start of 2020 until 24^th^ January 2021, when the last death from COVID-19 was recorded for this data download, there were 894 people who had died with a COVID-19 cause documented on their death certificates (ICD10 codes U07.1 and U07.2). To minimise potential biases, we used as controls people who died from other causes, but only during the same time interval, 4562 persons. The mean ages of the two groups were 74.1 (SD = 6.3) vs 75.0 (SD = 5.7) for the non-COVID-19 and COVID-19 deaths, respectively. This difference is small but significant at p = 6.3 × 10^−5^. For comparisons with people who had not died, we used 468,482 people who were still alive by March 2021 (i.e., we excluded those who had died prior to 2020). The mean age of people alive in March 2021 was much lower: 68.1 years (SD = 8.1).

The following characteristics were used as co-variates and were obtained at the time of recruitment when participants were seen at the assessment centres between 2006 and 2010: sex, body mass index (BMI), Townsend deprivation index (a measure of social deprivation) and ethnicity (divided into White British and Irish versus others, according to their self-report).

For medical comorbidities we used the latest available data on Category 1712, “First Occurrences” (downloaded on 7^th^ March 2021). These have been developed by the UKBB team using data from self-report, hospital admissions, general practitioners records and death certificates. They are divided according to the two-digit ICD10 codes. We chose only conditions found in at least 500 participants, in order to reduce the multiple testing problems, resulting in 334 ICD10 codes. As we wanted to analyse only pre-existing co-morbidities, we excluded events that first happened during 2020 which could have caused the death of the person, e.g. myocardial infarct, pulmonary embolism, stroke, renal or heart failure. Otherwise, such conditions could wrongly be identified as protective factors for a COVID-related death, being found more commonly among people dying from other causes.

### Statistical analysis

We tested the association between 334 disorders and COVID deaths against deaths from other causes, COVID deaths against those alive and non-COVID deaths against those alive. This was done using a logistic regression model including sex, age, BMI, Townsend deprivation index and ethnicity. This analysis was carried out in R, using the glm() function. Regression analyses were corrected for multiple testing using the Bonferroni correction method in the p.adjust() function in R.

## Results

There were 894 people who died from COVID-19 between the start of 2020 up to 24^th^ January 2021, another 4,562 who had died from other causes during the same time interval and 468,482 people who were still alive. We first tested the risks (ORs) of dying from COVID-19 vs. remaining alive for 29 ICD10 groups of psychiatric disorders. We used logistic regression with sex, age, BMI, Townsend deprivation index and ethnicity as co-variates (Figure 1).

**Figure 1.**
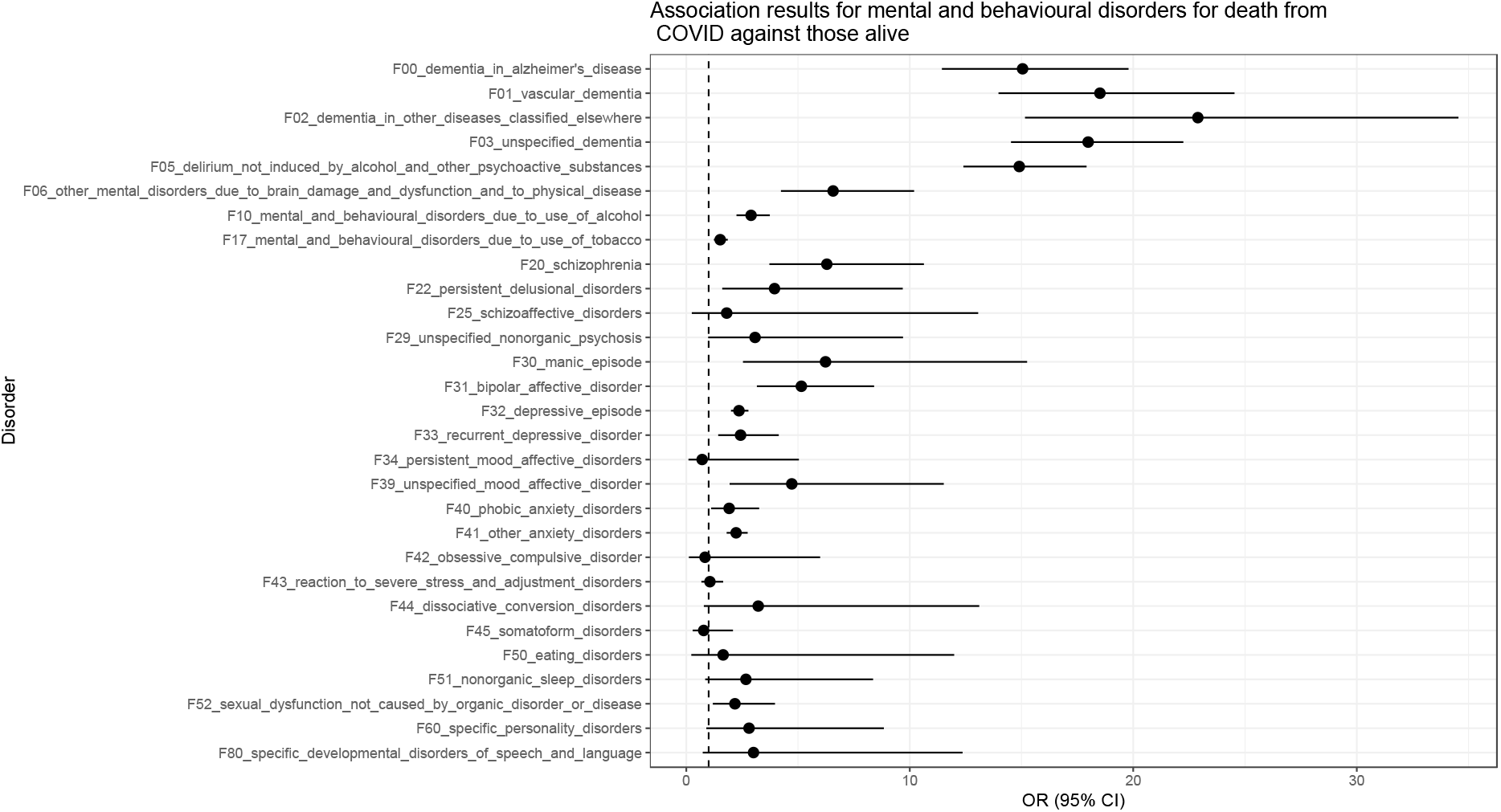
Association results for COVID death for people with mental and behavioural disorders against those still alive

The dementias stood out with ORs of around 10-20, implying a huge risk. However, people with dementia, and indeed with other severe psychiatric disorders, are at increased risk of dying from other causes, as explained in the Introduction. This was indeed the case for UKBB participants who had increased ORs for also dying from other disorders since the start of 2020 (Figure 2).

**Figure 2.**
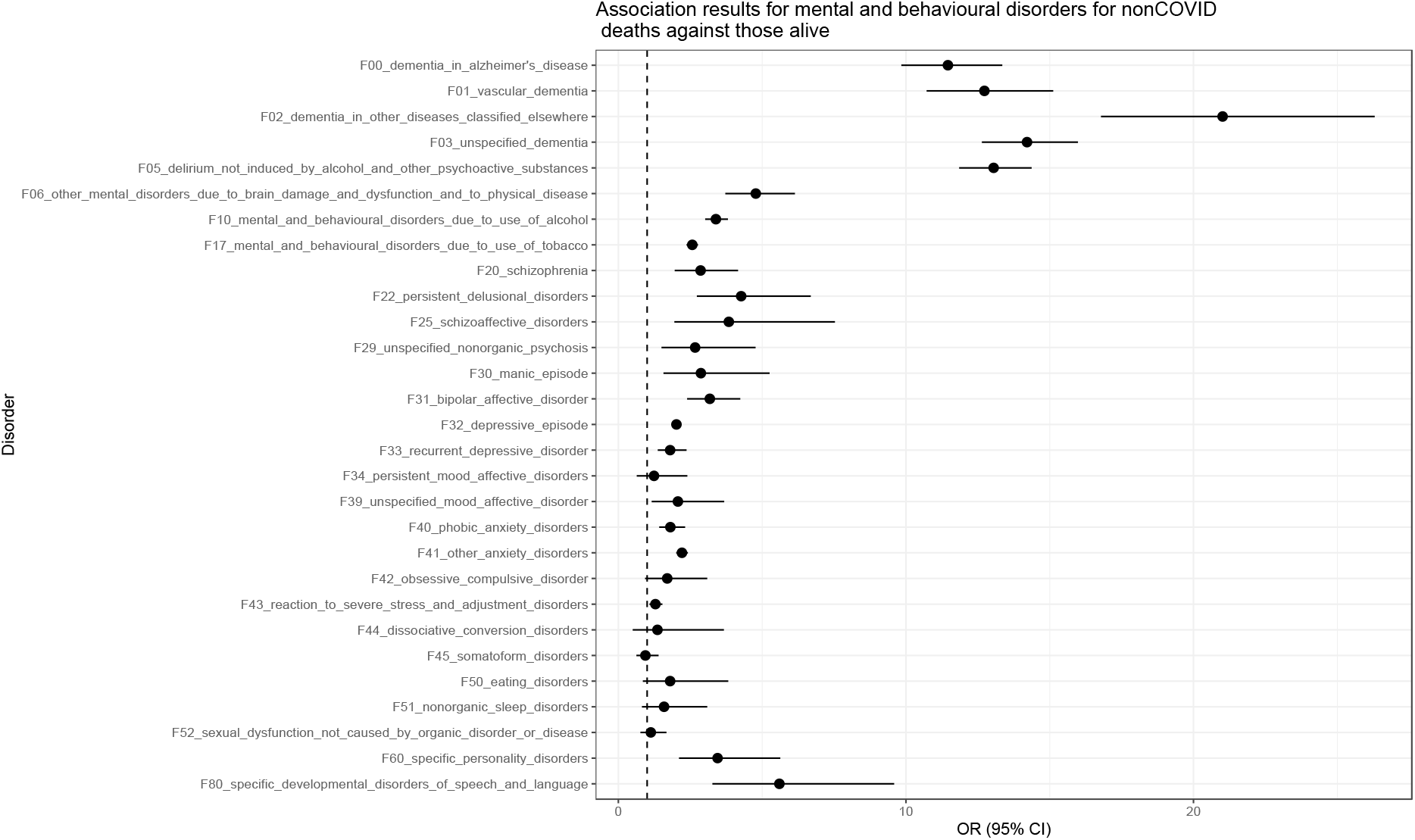
Association results for non-COVID death for people with mental and behavioural disorders, against those still alive

Both the risks for dying from COVID-19 and from other causes were increased and the patterns looked quite similar (Figures 1 & 2). We therefore examined if COVID-19 had any specificity in increasing risk over and above the other general causes for death. We compared the risk for dying from COVID-19 vs. dying from other causes since the start of 2020, using logistic regression with the same co-variates (Figure 3).

**Figure 3.**
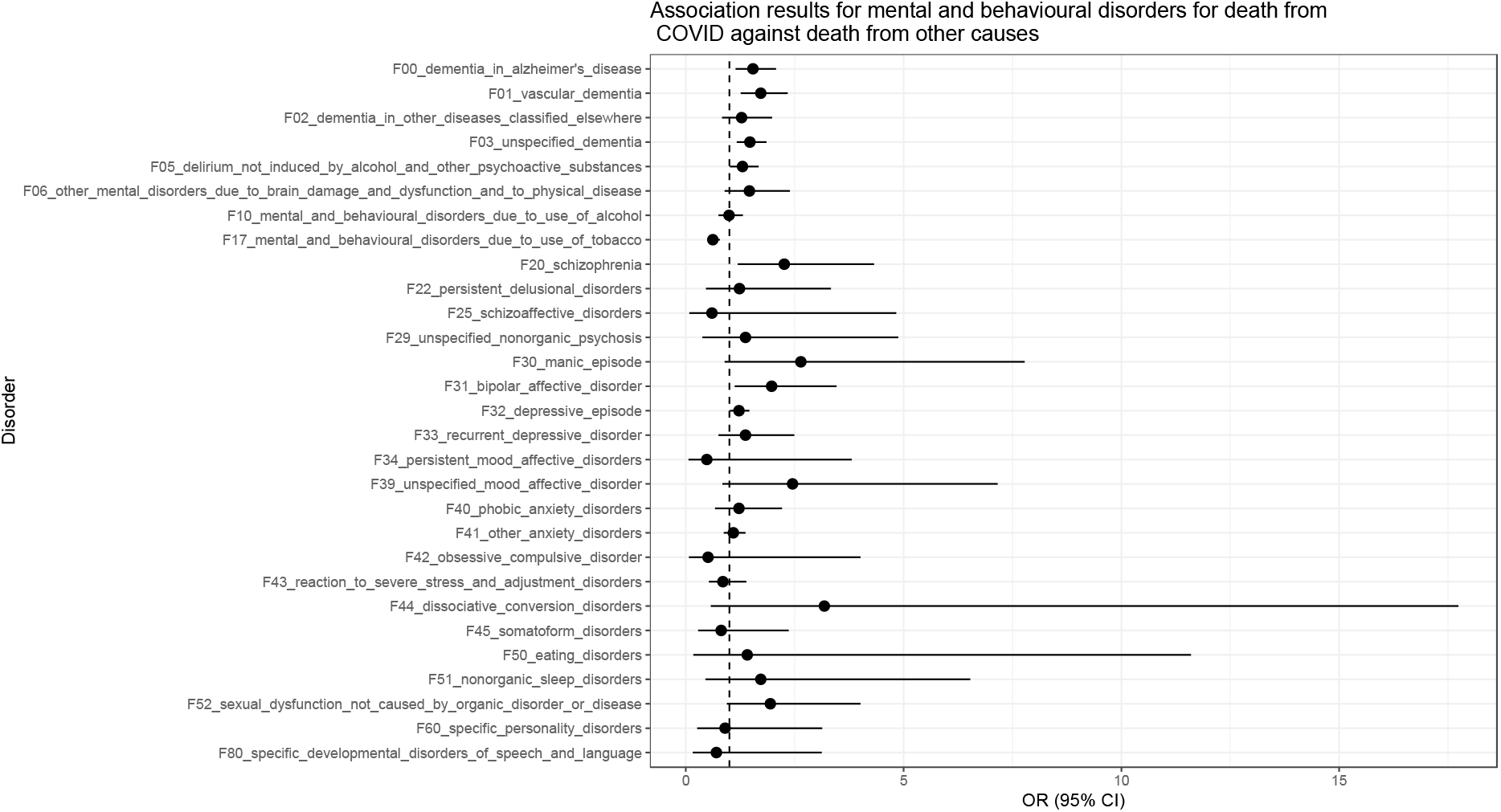
Association results for death from COVID against death from other causes for mental and behavioural disorders

The ORs were substantially reduced but eight disorders remained significant at p < 0.05, seven of them in the direction of increasing risk: Alzheimer’s disease, vascular and unspecified dementia, schizophrenia, bipolar disorder, depressive episodes and delirium. Mental disorders due to the use of tobacco (i.e. smoking addiction) were also significant but as a protective factor (see Discussion). In order to compare the mental health disorders with other medical comorbidities (the remaining 305 ICD10 codes), we present all nominally significant results in Figure 4. All comparisons between COVID-19 and deaths due to other causes are available in Supplementary Table 1.

**Table 1.**
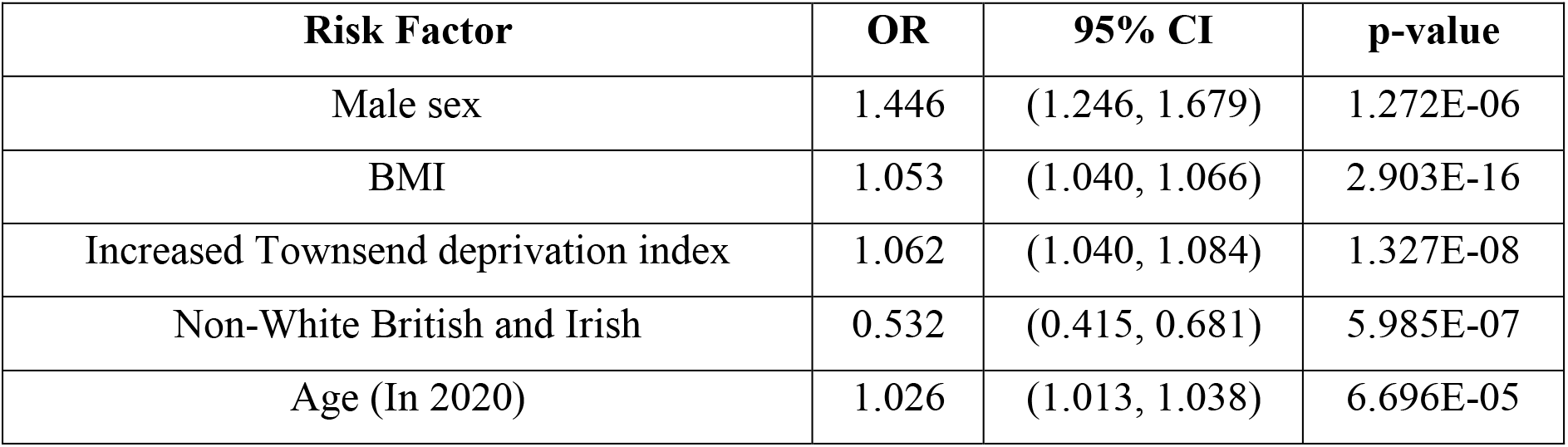
Risk factors associated with COVID death when compared to death from other causes.

**Figure 4.**
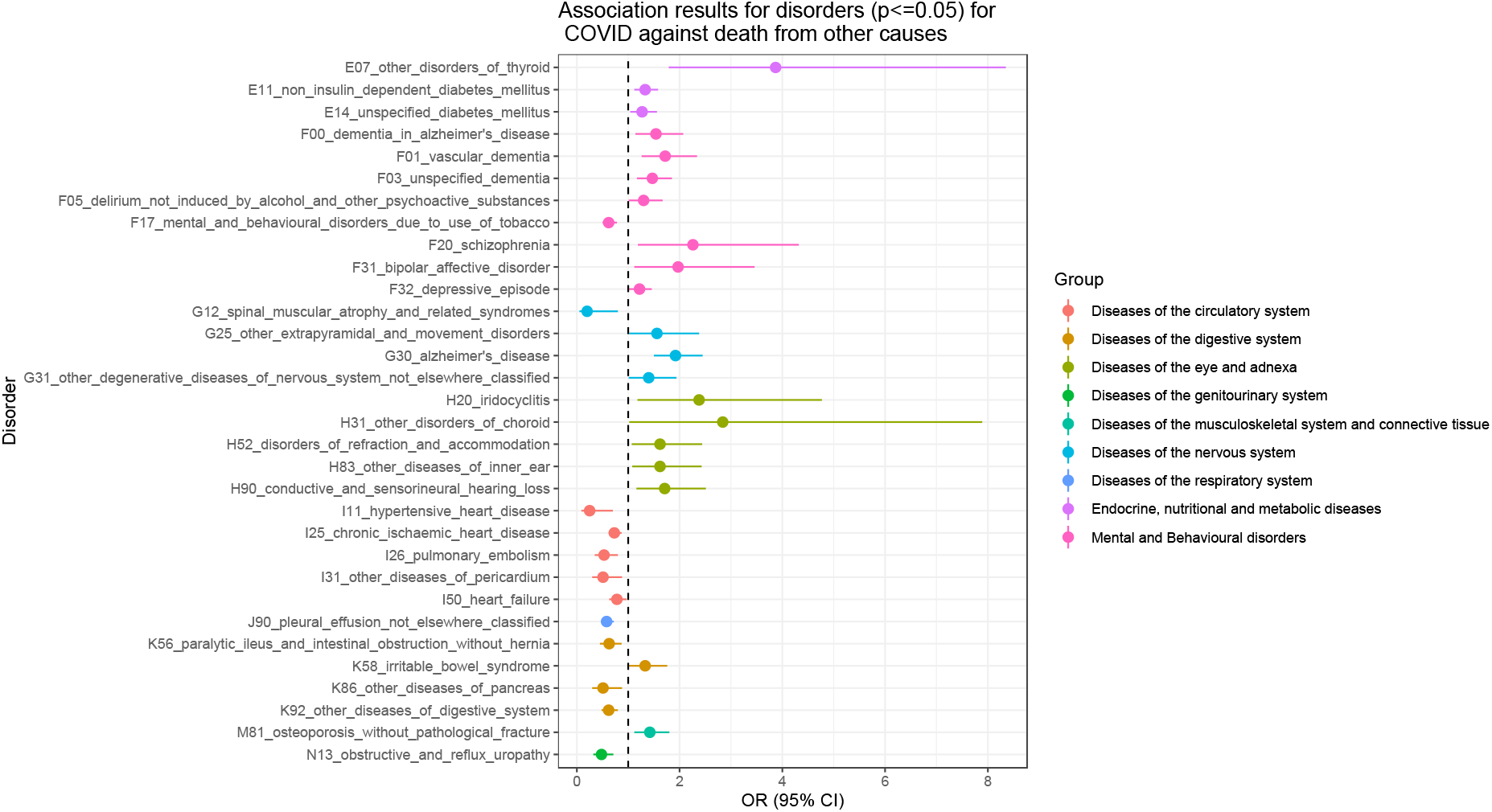
Nominally significant (p<=0.05) association results for death from COVID against death from other causes for any medical co-morbid conditions

Age, sex, social deprivation, BMI and ethnicity were significantly associated with increased risk for COVID death when compared to death from other causes, in line with previous findings (Table 1).

## Discussion

Individuals with schizophrenia and bipolar disorder have a reduced life expectancy of 15-20 years compared to the general population (Lawrence et al, 2013) with cardiovascular disease, COPD, accidental death, influenza and pneumonia contributing most of the increased mortality (Olfson et al, 2015; Nielsen et al, 2021). Such people are also more likely to die from COVID-19 (Toubasi et al, 2021). This raises the question whether the increased rate of COVID-19 deaths among patients with severe psychiatric disorders is not simply a reflection of their increased risk of dying prematurely, and that COVID-19 might have the same effect on mortality regardless of any mental health problems. We demonstrate that in such patients the risk for a COVID-19 death is over and above the risk for dying from other causes. To illustrate the point, the OR for dying from COVID for patients with Alzheimer’s disease (F00) was 15.0 (95% CI = 11.4 - 19.8) compared to those who stayed alive. However, people with this diagnosis also died at high rates from other causes: OR = 11.5 (95% CI = 9.8 – 13.4). The risk was still significantly increased when the two causes for death were compared, but with a much reduced effect: OR = 1.5 (95% CI = 1.1 – 1.9). Similarly, for schizophrenia (F20) the OR for dying from COVID-19 was 6.3 (95% CI = 3.7 – 10.6) and although going down, it remained significantly increased, compared to patients with schizophrenia who died from other causes: OR = 2.3 (95% CI = 1.2 – 4.3). There was a trend for the degree of risk to correlate with the severity of the psychiatric condition: schizophrenia had the highest risk with OR = 2.3, followed by bipolar disorder (OR = 2.0) and the risk was much lower for depression (OR = 1.2).

Using this approach, we observe that mental health conditions appear to be over-represented among those dying from COVID-19, compared to any other group of medical co-morbidities, accounting for seven out of only 20 nominally significant associations which increase risk (Figure 4). A number of medical conditions appeared to act as “protective” factors, as they lead to premature death in their own right, without the added risk from COVID-19 infection. Of the previously implicated medical co-morbidities, only diabetes was associated with specifically increased risk for COVID-19 death, while hypertension, chronic ischaemic heart disease and heart failure were associated with reduced risk, and chronic obstructive pulmonary disease (COPD, J43: unspecified chronic bronchitis) was not significantly associated.

The question arises whether patients with psychiatric disorders are at a higher risk because of their known medical co-morbidities and socio-demographic background, which happen to also be risk factors for dying form COVID-19, such as diabetes, COPD, increased BMI and social deprivation. Increased BMI and social deprivation are over-represented among people with mental health problems, due to changes in lifestyle, loss of earnings or effects of the medication. There was indeed a specificity for COVID-19 death for the known associations with increased BMI, male sex, social deprivation and ethnicity (Table 1). However, these were already used as a co-variates in all analyses. We also examined whether the effect from schizophrenia (the condition with the highest OR) was mediated by the increased rate of diabetes, ischaemic heart disease, myocardial infarction and COPD among such patients. There was no significant interaction between schizophrenia and these conditions, possibly due to the small sample size (data not presented). About half of the patients with schizophrenia who died from COVID-19 had one of these four co-morbidities (eight out of 15, 53.3%), but similarly, 19 out of 32 patients with schizophrenia who died from other causes, also had one of these comorbidities (59.4%). In contrast, only 14.4% of participants who stayed alive had one of these four comorbidities.

There must be other factors that increase susceptibility to dying from COVID-19 infections among people with mental health problems. For the dementias, it is possible that the infections spreading in nursing homes during the first wave of the pandemic in the UK, have increased the exposure of such patients to the virus. For schizophrenia, depression and bipolar disorder it is more difficult to understand what factors beyond medical comorbidities contribute directly to increased mortality. Differences in lifestyle, reduced exercise, stress and the effects of medication might be reducing the immunity or resilience to illness and infection, or such people might be more reluctant to seek medical help and delay their treatment. We are unable to speculate further based on this analysis.

The other disorders which appeared over-represented were the sensory conditions: conductive and sensorineural hearing loss, other diseases of inner ear, refraction and accommodation disorders, iridocyclitis. We are unable to hazard any explanation for these associations, which could also reflect some biases. Among the medical co-morbidities we do observe modestly increased odds ratios for diabetes. However, hypertension, cardiac or respiratory disorders did not incur an increase in specific risk. In fact, several of these conditions showed a reduced risk, indicating that such patients were more likely to die from other causes, including the disorders in question, or might have been shielding during the pandemic. Thus, history of pleural effusion, chronic ischaemic heart disease, pulmonary embolism, paralytic ileus and several other serious medical conditions were associated with significant risk of dying with a COVID-19 infection. Another interesting observation is the lowered risk among those addicted to smoking (F17), which is one of only three associations that would survive correction for multiple testing, and thus should not be dismissed as a false-positive finding. This association has been reported before and has been subjected to a lot of analysis. It is believed to be due to biases and that it is unlikely to lower the risk of death. In the current analysis it could be explained by the increased risk of dying from other conditions among smokers, similar to the association with ischaemic heart disease, which also appears to lower the risk for dying from COVID, but only reflects the high risk of dying as a consequence of the heart disease.

In conclusion, we show that psychiatric patients appear to have an increased risk for dying from COVID-19, although we are unable to identify the specific reasons behind this association. Such patients should receive special attention during this pandemic, including being prioritised for vaccination, like those with serious medical conditions.

## Data Availability

All data are available via the UK Biobank resourse

## Acknowledgements

This research has been conducted using the UK Biobank Resource under Application Number 14421 and COVID-19. The work at Cardiff University was funded by the Medical Research Council (MRC) Centre Grant (MR/L010305/1).

