## Supplemental Table 1 for "COVID-19 and mortality risk in patients with psychiatric disorders"

**COVID-19 and risk of death among patients with psychiatric disorders**

**Supplementary Table 1**

| **Disorder** | **Ncases** | **Ncontrols** | **OR** | **95% CI** | **P** | **Padj** |
| --- | --- | --- | --- | --- | --- | --- |
| G30_alzheimer's_disease | 412 | 5044 | 1.92 | (1.5-2.45) | 2.28E-07 | 7.86E-05 |
| J90_pleural_effusion_not_elsewhere_classified | 1010 | 4446 | 0.58 | (0.47-0.72) | 6.73E-07 | 0.00023 |
| F17_mental_and_behavioural_disorders_due_to_use_of_tobacco | 955 | 4501 | 0.62 | (0.5-0.78) | 2.43E-05 | 0.00835 |
| K92_other_diseases_of_digestive_system | 682 | 4774 | 0.62 | (0.48-0.8) | 0.00018 | 0.06086 |
| I25_chronic_ischaemic_heart_disease | 1680 | 3776 | 0.73 | (0.62-0.87) | 0.00027 | 0.09417 |
| N13_obstructive_and_reflux_uropathy | 324 | 5132 | 0.48 | (0.32-0.71) | 0.00028 | 0.09747 |
| F01_vascular_dementia | 244 | 5212 | 1.72 | (1.26-2.34) | 0.00055 | 0.18831 |
| E07_other_disorders_of_thyroid | 30 | 5426 | 3.87 | (1.79-8.35) | 0.00057 | 0.19740 |
| F03_unspecified_dementia | 529 | 4927 | 1.47 | (1.17-1.85) | 0.00106 | 0.36391 |
| E11_non_insulin_dependent_diabetes_mellitus | 1312 | 4144 | 1.33 | (1.12-1.58) | 0.00113 | 0.38981 |
| I26_pulmonary_embolism | 282 | 5174 | 0.53 | (0.35-0.8) | 0.00252 | 0.86603 |
| M81_osteoporosis_without_pathological_fracture | 607 | 4849 | 1.42 | (1.12-1.8) | 0.00364 | 1 |
| F00_dementia_in_alzheimer's_disease | 286 | 5170 | 1.54 | (1.14-2.07) | 0.00467 | 1 |
| K56_paralytic_ileus_and_intestinal_obstruction_without_hernia | 420 | 5036 | 0.63 | (0.45-0.87) | 0.00478 | 1 |
| H90_conductive_and_sensorineural_hearing_loss | 145 | 5311 | 1.71 | (1.16-2.51) | 0.00622 | 1 |
| I11_hypertensive_heart_disease | 67 | 5389 | 0.25 | (0.09-0.7) | 0.00814 | 1 |
| F20_schizophrenia | 47 | 5409 | 2.26 | (1.19-4.32) | 0.01304 | 1 |
| H20_iridocyclitis | 38 | 5418 | 2.38 | (1.18-4.77) | 0.01482 | 1 |
| K86_other_diseases_of_pancreas | 171 | 5285 | 0.51 | (0.3-0.88) | 0.01544 | 1 |
| I31_other_diseases_of_pericardium | 160 | 5296 | 0.51 | (0.3-0.88) | 0.01573 | 1 |
| F31_bipolar_affective_disorder | 67 | 5389 | 1.97 | (1.12-3.46) | 0.01840 | 1 |
| H83_other_diseases_of_inner_ear | 138 | 5318 | 1.62 | (1.08-2.43) | 0.01928 | 1 |
| E14_unspecified_diabetes_mellitus | 747 | 4709 | 1.27 | (1.04-1.56) | 0.01970 | 1 |
| H52_disorders_of_refraction_and_accommodation | 135 | 5321 | 1.62 | (1.07-2.44) | 0.02117 | 1 |
| G12_spinal_muscular_atrophy_and_related_syndromes | 62 | 5394 | 0.2 | (0.05-0.8) | 0.02379 | 1 |
| I50_heart_failure | 767 | 4689 | 0.78 | (0.63-0.97) | 0.02454 | 1 |
| F32_depressive_episode | 1051 | 4405 | 1.22 | (1.01-1.46) | 0.03754 | 1 |
| G25_other_extrapyramidal_and_movement_disorders | 136 | 5320 | 1.56 | (1.02-2.38) | 0.04062 | 1 |
| G31_other_degenerative_diseases_of_nervous_system_not_elsewhere_classified | 257 | 5199 | 1.4 | (1.01-1.94) | 0.04117 | 1 |
| F05_delirium_not_induced_by_alcohol_and_other_psychoactive_substances | 453 | 5003 | 1.3 | (1.01-1.67) | 0.04130 | 1 |
| H31_other_disorders_of_choroid | 18 | 5438 | 2.84 | (1.02-7.89) | 0.04551 | 1 |
| K58_irritable_bowel_syndrome | 383 | 5073 | 1.33 | (1.01-1.76) | 0.04555 | 1 |
| I83_varicose_veins_of_lower_extremities | 404 | 5052 | 0.75 | (0.55-1.01) | 0.05414 | 1 |
| D64_other_anaemias | 1290 | 4166 | 0.84 | (0.7-1.01) | 0.05773 | 1 |
| M19_other_arthrosis | 1649 | 3807 | 1.16 | (0.99-1.37) | 0.06373 | 1 |
| I85_oesophageal_varices | 85 | 5371 | 0.52 | (0.26-1.05) | 0.06722 | 1 |
| H60_otitis_externa | 292 | 5164 | 1.32 | (0.98-1.78) | 0.06926 | 1 |
| F52_sexual_dysfunction_not_caused_by_organic_disorder_or_disease | 38 | 5418 | 1.94 | (0.94-4.01) | 0.07116 | 1 |
| G70_myasthenia_gravis_and_other_myoneural_disorders | 15 | 5441 | 2.66 | (0.9-7.82) | 0.07633 | 1 |
| F30_manic_episode | 16 | 5440 | 2.64 | (0.89-7.78) | 0.07905 | 1 |
| H49_paralytic_strabismus | 37 | 5419 | 0.28 | (0.07-1.17) | 0.08020 | 1 |
| H92_otalgia_and_effusion_of_ear | 183 | 5273 | 1.39 | (0.96-2.03) | 0.08349 | 1 |
| L91_hypertrophic_disorders_of_skin | 75 | 5381 | 0.5 | (0.23-1.1) | 0.08424 | 1 |
| N95_menopausal_and_other_perimenopausal_disorders | 353 | 5103 | 1.32 | (0.96-1.81) | 0.08433 | 1 |
| K70_alcoholic_liver_disease | 149 | 5307 | 0.63 | (0.38-1.07) | 0.08600 | 1 |
| K81_cholecystitis | 171 | 5285 | 1.38 | (0.95-2) | 0.09419 | 1 |
| I44_atrioventricular_and_left_bundle_branch_block | 471 | 4985 | 0.8 | (0.61-1.04) | 0.09895 | 1 |
| D56_thalassaemia | 5 | 5451 | 4.65 | (0.74-29.3) | 0.10203 | 1 |
| F39_unspecified_mood_affective_disorder | 17 | 5439 | 2.45 | (0.84-7.16) | 0.10241 | 1 |
| N04_nephrotic_syndrome | 26 | 5430 | 2.06 | (0.85-4.98) | 0.10756 | 1 |
| G51_facial_nerve_disorders | 61 | 5395 | 0.5 | (0.21-1.17) | 0.10977 | 1 |
| I49_other_cardiac_arrhythmias | 234 | 5222 | 0.73 | (0.5-1.07) | 0.11129 | 1 |
| H93_other_disorders_of_ear_not_elsewhere_classified | 200 | 5256 | 1.33 | (0.93-1.91) | 0.11357 | 1 |
| K40_inguinal_hernia | 521 | 4935 | 0.81 | (0.62-1.06) | 0.11927 | 1 |
| H66_suppurative_and_unspecified_otitis_media | 113 | 5343 | 1.43 | (0.9-2.27) | 0.12524 | 1 |
| N35_urethral_stricture | 120 | 5336 | 0.66 | (0.38-1.13) | 0.13108 | 1 |
| F06_other_mental_disorders_due_to_brain_damage_and_dysfunction_and_to_physical_disease | 92 | 5364 | 1.46 | (0.89-2.39) | 0.13510 | 1 |
| K35_acute_appendicitis | 55 | 5401 | 0.49 | (0.19-1.25) | 0.13554 | 1 |
| K83_other_diseases_of_biliary_tract | 275 | 5181 | 0.75 | (0.52-1.09) | 0.13691 | 1 |
| N80_endometriosis | 61 | 5395 | 0.46 | (0.16-1.28) | 0.13724 | 1 |
| N97_female_infertility | 18 | 5438 | 2.37 | (0.75-7.52) | 0.14293 | 1 |
| I67_other_cerebrovascular_diseases | 623 | 4833 | 1.18 | (0.94-1.47) | 0.14666 | 1 |
| J93_pneumothorax | 111 | 5345 | 1.42 | (0.88-2.3) | 0.14977 | 1 |
| N39_other_disorders_of_urinary_system | 1627 | 3829 | 1.12 | (0.96-1.32) | 0.15420 | 1 |
| K43_ventral_hernia | 173 | 5283 | 0.72 | (0.46-1.13) | 0.15755 | 1 |
| M35_other_systemic_involvement_of_connective_tissue | 173 | 5283 | 1.32 | (0.89-1.96) | 0.16051 | 1 |
| K64_haemorrhoids_and_perianal_venous_thrombosis | 571 | 4885 | 0.84 | (0.66-1.08) | 0.17493 | 1 |
| H05_disorders_of_orbit | 23 | 5433 | 0.25 | (0.03-1.9) | 0.18062 | 1 |
| E78_disorders_of_lipoprotein_metabolism_and_other_lipidaemias | 2346 | 3110 | 1.11 | (0.95-1.29) | 0.18129 | 1 |
| M60_myositis | 20 | 5436 | 0.25 | (0.03-1.91) | 0.18181 | 1 |
| G37_other_demyelinating_diseases_of_central_nervous_system | 8 | 5448 | 3.22 | (0.57-18.06) | 0.18349 | 1 |
| H35_other_retinal_disorders | 338 | 5118 | 1.21 | (0.91-1.61) | 0.18375 | 1 |
| I84_haemorrhoids | 412 | 5044 | 0.82 | (0.62-1.1) | 0.18591 | 1 |
| E66_obesity | 958 | 4498 | 1.15 | (0.94-1.41) | 0.18614 | 1 |
| F44_dissociative_conversion_disorders | 7 | 5449 | 3.18 | (0.57-17.74) | 0.18716 | 1 |
| H36_retinal_disorders_in_diseases_classified_elsewhere | 182 | 5274 | 1.28 | (0.89-1.84) | 0.18866 | 1 |
| I22_subsequent_myocardial_infarction | 42 | 5414 | 0.53 | (0.21-1.38) | 0.19414 | 1 |
| M17_gonarthrosis_arthrosis_of_knee | 678 | 4778 | 0.86 | (0.69-1.08) | 0.19615 | 1 |
| N19_unspecified_renal_failure | 149 | 5307 | 0.73 | (0.45-1.18) | 0.19894 | 1 |
| K55_vascular_disorders_of_intestine | 144 | 5312 | 0.72 | (0.44-1.19) | 0.20164 | 1 |
| E22_hyperfunction_of_pituitary_gland | 58 | 5398 | 0.55 | (0.22-1.39) | 0.20302 | 1 |
| N70_salpingitis_and_oophoritis | 6 | 5450 | 3.06 | (0.54-17.19) | 0.20448 | 1 |
| K75_other_inflammatory_liver_diseases | 99 | 5357 | 0.67 | (0.36-1.24) | 0.20459 | 1 |
| H65_nonsuppurative_otitis_media | 68 | 5388 | 1.46 | (0.81-2.63) | 0.21298 | 1 |
| I21_acute_myocardial_infarction | 624 | 4832 | 0.86 | (0.68-1.09) | 0.21332 | 1 |
| E83_disorders_of_mineral_metabolism | 617 | 4839 | 0.86 | (0.67-1.09) | 0.21658 | 1 |
| H01_other_inflammation_of_eyelid | 91 | 5365 | 0.67 | (0.35-1.27) | 0.21724 | 1 |
| I38_endocarditis_valve_unspecified | 75 | 5381 | 0.66 | (0.33-1.3) | 0.22579 | 1 |
| H10_conjunctivitis | 293 | 5163 | 1.21 | (0.89-1.64) | 0.22608 | 1 |
| M43_other_deforming_dorsopathies | 139 | 5317 | 0.73 | (0.44-1.21) | 0.22679 | 1 |
| K82_other_diseases_of_gallbladder | 84 | 5372 | 0.65 | (0.32-1.32) | 0.23150 | 1 |
| H81_disorders_of_vestibular_function | 160 | 5296 | 1.28 | (0.85-1.92) | 0.23390 | 1 |
| H04_disorders_of_lachrymal_system | 251 | 5205 | 0.8 | (0.55-1.16) | 0.23805 | 1 |
| D57_sickle_cell_disorders | 10 | 5446 | 2.18 | (0.59-8.08) | 0.24456 | 1 |
| I62_other_nontraumatic_intracranial_haemorrhage | 38 | 5418 | 0.54 | (0.19-1.54) | 0.24748 | 1 |
| H61_other_disorders_of_external_ear | 441 | 5015 | 0.85 | (0.64-1.12) | 0.24868 | 1 |
| N21_calculus_of_lower_urinary_tract | 61 | 5395 | 0.64 | (0.3-1.37) | 0.25233 | 1 |
| F02_dementia_in_other_diseases_classified_elsewhere | 142 | 5314 | 1.28 | (0.83-1.98) | 0.26112 | 1 |
| I65_occlusion_and_stenosis_of_precerebral_arteries_not_resulting_in_cerebral_infarction | 124 | 5332 | 0.74 | (0.44-1.25) | 0.26351 | 1 |
| N43_hydrocele_and_spermatocele | 61 | 5395 | 1.4 | (0.77-2.55) | 0.27425 | 1 |
| M05_seropositive_rheumatoid_arthritis | 45 | 5411 | 1.49 | (0.73-3.08) | 0.27593 | 1 |
| E27_other_disorders_of_adrenal_gland | 81 | 5375 | 1.37 | (0.78-2.4) | 0.27653 | 1 |
| D52_folate_deficiency_anaemia | 65 | 5391 | 1.4 | (0.76-2.58) | 0.28113 | 1 |
| D50_iron_deficiency_anaemia | 828 | 4628 | 0.89 | (0.73-1.1) | 0.28259 | 1 |
| H02_other_disorders_of_eyelid | 273 | 5183 | 1.19 | (0.87-1.63) | 0.28537 | 1 |
| M10_gout | 500 | 4956 | 1.14 | (0.9-1.44) | 0.28670 | 1 |
| H43_disorders_of_vitreous_body | 164 | 5292 | 0.78 | (0.49-1.24) | 0.28916 | 1 |
| H34_retinal_vascular_occlusions | 60 | 5396 | 1.4 | (0.75-2.62) | 0.29381 | 1 |
| J42_unspecified_chronic_bronchitis | 29 | 5427 | 0.52 | (0.16-1.76) | 0.29397 | 1 |
| H54_blindness_and_low_vision | 147 | 5309 | 0.78 | (0.48-1.25) | 0.29505 | 1 |
| J84_other_interstitial_pulmonary_diseases | 307 | 5149 | 1.17 | (0.87-1.59) | 0.29683 | 1 |
| I35_nonrheumatic_aortic_valve_disorders | 361 | 5095 | 0.86 | (0.64-1.15) | 0.30092 | 1 |
| H68_eustachian_salpingitis_and_obstruction | 28 | 5428 | 1.59 | (0.66-3.8) | 0.30100 | 1 |
| J95_postprocedural_respiratory_disorders_not_elsewhere_classified | 41 | 5415 | 0.58 | (0.2-1.64) | 0.30313 | 1 |
| L94_other_localised_connective_tissue_disorders | 15 | 5441 | 0.34 | (0.04-2.66) | 0.30429 | 1 |
| N85_other_noninflammatory_disorders_of_uterus_except_cervix | 77 | 5379 | 0.66 | (0.3-1.46) | 0.30510 | 1 |
| F33_recurrent_depressive_disorder | 68 | 5388 | 1.37 | (0.75-2.49) | 0.30917 | 1 |
| M11_other_crystal_arthropathies | 29 | 5427 | 1.56 | (0.66-3.73) | 0.31308 | 1 |
| N92_excessive_frequent_and_irregular_menstruation | 151 | 5305 | 1.27 | (0.79-2.04) | 0.31782 | 1 |
| H47_other_disorders_of_optic_2nd_nerve_and_visual_pathways | 40 | 5416 | 0.59 | (0.21-1.67) | 0.31813 | 1 |
| J44_other_chronic_obstructive_pulmonary_disease | 1014 | 4442 | 1.1 | (0.91-1.32) | 0.32209 | 1 |
| K51_ulcerative_colitis | 123 | 5333 | 0.76 | (0.44-1.32) | 0.32562 | 1 |
| N41_inflammatory_diseases_of_prostate | 78 | 5378 | 1.32 | (0.76-2.29) | 0.32625 | 1 |
| I64_stroke_not_specified_as_haemorrhage_or_infarction | 278 | 5178 | 1.17 | (0.86-1.59) | 0.32945 | 1 |
| M80_osteoporosis_with_pathological_fracture | 109 | 5347 | 1.28 | (0.78-2.12) | 0.33043 | 1 |
| K57_diverticular_disease_of_intestine | 1202 | 4254 | 0.92 | (0.77-1.1) | 0.33436 | 1 |
| K30_dyspepsia | 243 | 5213 | 1.18 | (0.84-1.67) | 0.33560 | 1 |
| N02_recurrent_and_persistent_haematuria | 47 | 5409 | 0.65 | (0.28-1.55) | 0.33670 | 1 |
| I82_other_venous_embolism_and_thrombosis | 74 | 5382 | 0.69 | (0.33-1.47) | 0.33889 | 1 |
| N50_other_disorders_of_male_genital_organs | 189 | 5267 | 1.2 | (0.83-1.73) | 0.34227 | 1 |
| I46_cardiac_arrest | 58 | 5398 | 0.69 | (0.32-1.48) | 0.34422 | 1 |
| K20_oesophagitis | 351 | 5105 | 1.15 | (0.86-1.53) | 0.34832 | 1 |
| I70_atherosclerosis | 216 | 5240 | 0.83 | (0.56-1.23) | 0.34837 | 1 |
| G41_status_epilepticus | 27 | 5429 | 0.5 | (0.11-2.15) | 0.34862 | 1 |
| E80_disorders_of_porphyrin_and_bilirubin_metabolism | 32 | 5424 | 0.57 | (0.17-1.88) | 0.35266 | 1 |
| H33_retinal_detachments_and_breaks | 83 | 5373 | 0.74 | (0.39-1.41) | 0.35378 | 1 |
| I86_varicose_veins_of_other_sites | 54 | 5402 | 0.69 | (0.3-1.55) | 0.36321 | 1 |
| H16_keratitis | 34 | 5422 | 0.62 | (0.21-1.78) | 0.37184 | 1 |
| H71_cholesteatoma_of_middle_ear | 13 | 5443 | 0.39 | (0.05-3.06) | 0.37218 | 1 |
| G20_parkinson's_disease | 313 | 5143 | 1.15 | (0.85-1.56) | 0.37255 | 1 |
| I77_other_disorders_of_arteries_and_arterioles | 155 | 5301 | 0.81 | (0.51-1.29) | 0.37313 | 1 |
| I10_essential_primary_hypertension | 3414 | 2042 | 1.08 | (0.91-1.27) | 0.37425 | 1 |
| G81_hemiplegia | 245 | 5211 | 1.17 | (0.83-1.65) | 0.37626 | 1 |
| I80_phlebitis_and_thrombophlebitis | 495 | 4961 | 0.89 | (0.68-1.16) | 0.38357 | 1 |
| I61_intracerebral_haemorrhage | 61 | 5395 | 1.33 | (0.69-2.54) | 0.39121 | 1 |
| K50_crohn's_disease_regional_enteritis | 72 | 5384 | 0.72 | (0.34-1.52) | 0.39241 | 1 |
| E21_hyperparathyroidism_and_other_disorders_of_parathyroid_gland | 72 | 5384 | 1.3 | (0.71-2.37) | 0.39322 | 1 |
| N48_other_disorders_of_penis | 163 | 5293 | 1.19 | (0.8-1.75) | 0.39342 | 1 |
| K59_other_functional_intestinal_disorders | 1368 | 4088 | 0.93 | (0.78-1.1) | 0.39438 | 1 |
| M06_other_rheumatoid_arthritis | 337 | 5119 | 1.13 | (0.85-1.52) | 0.39896 | 1 |
| H25_senile_cataract | 570 | 4886 | 0.9 | (0.71-1.15) | 0.40422 | 1 |
| N28_other_disorders_of_kidney_and_ureter_not_elsewhere_classified | 355 | 5101 | 0.88 | (0.65-1.19) | 0.40438 | 1 |
| L29_pruritus | 229 | 5227 | 1.16 | (0.82-1.65) | 0.40567 | 1 |
| G50_disorders_of_trigeminal_nerve | 52 | 5404 | 0.7 | (0.29-1.65) | 0.41060 | 1 |
| M15_polyarthrosis | 275 | 5181 | 0.87 | (0.62-1.22) | 0.41640 | 1 |
| N46_male_infertility | 8 | 5448 | 1.95 | (0.39-9.92) | 0.41866 | 1 |
| H50_other_strabismus | 44 | 5412 | 0.65 | (0.22-1.87) | 0.41936 | 1 |
| K29_gastritis_and_duodenitis | 1087 | 4369 | 1.08 | (0.9-1.29) | 0.41988 | 1 |
| M41_scoliosis | 106 | 5350 | 1.24 | (0.73-2.12) | 0.42713 | 1 |
| F51_nonorganic_sleep_disorders | 13 | 5443 | 1.72 | (0.45-6.53) | 0.42763 | 1 |
| I34_nonrheumatic_mitral_valve_disorders | 259 | 5197 | 0.87 | (0.61-1.24) | 0.43498 | 1 |
| N63_unspecified_lump_in_breast | 130 | 5326 | 1.22 | (0.74-2.01) | 0.43746 | 1 |
| M54_dorsalgia | 1282 | 4174 | 1.07 | (0.9-1.27) | 0.44271 | 1 |
| L90_atrophic_disorders_of_skin | 78 | 5378 | 0.77 | (0.39-1.52) | 0.44793 | 1 |
| H69_other_disorders_of_eustachian_tube | 18 | 5438 | 1.55 | (0.5-4.81) | 0.44856 | 1 |
| K25_gastric_ulcer | 287 | 5169 | 1.13 | (0.82-1.54) | 0.45449 | 1 |
| K31_other_diseases_of_stomach_and_duodenum | 441 | 5015 | 0.9 | (0.68-1.19) | 0.45801 | 1 |
| M50_cervical_disk_disorders | 95 | 5361 | 1.22 | (0.72-2.06) | 0.46057 | 1 |
| F41_other_anxiety_disorders | 639 | 4817 | 1.09 | (0.87-1.37) | 0.46336 | 1 |
| I33_acute_and_subacute_endocarditis | 43 | 5413 | 0.71 | (0.28-1.81) | 0.46808 | 1 |
| M47_spondylosis | 708 | 4748 | 1.08 | (0.87-1.34) | 0.47086 | 1 |
| N83_noninflammatory_disorders_of_ovary_fallopian_tube_and_broad_ligament | 116 | 5340 | 0.8 | (0.44-1.46) | 0.47429 | 1 |
| H18_other_disorders_of_cornea | 68 | 5388 | 0.77 | (0.38-1.58) | 0.47900 | 1 |
| L93_lupus_erythematosus | 13 | 5443 | 0.48 | (0.06-3.75) | 0.47996 | 1 |
| E06_thyroiditis | 13 | 5443 | 1.6 | (0.43-5.9) | 0.48363 | 1 |
| F34_persistent_mood_affective_disorders | 10 | 5446 | 0.48 | (0.06-3.81) | 0.48506 | 1 |
| I20_angina_pectoris | 988 | 4468 | 0.93 | (0.77-1.13) | 0.48656 | 1 |
| G82_paraplegia_and_tetraplegia | 32 | 5424 | 1.43 | (0.52-3.96) | 0.49039 | 1 |
| I87_other_disorders_of_veins | 82 | 5374 | 0.81 | (0.44-1.49) | 0.49236 | 1 |
| K61_abscess_of_anal_and_rectal_regions | 67 | 5389 | 0.78 | (0.38-1.6) | 0.49393 | 1 |
| E79_disorders_of_purine_and_pyrimidine_metabolism | 9 | 5447 | 0.48 | (0.06-3.97) | 0.49783 | 1 |
| L40_psoriasis | 254 | 5202 | 1.12 | (0.8-1.58) | 0.50189 | 1 |
| N93_other_abnormal_uterine_and_vaginal_bleeding | 77 | 5379 | 1.24 | (0.65-2.35) | 0.50985 | 1 |
| I73_other_peripheral_vascular_diseases | 485 | 4971 | 0.92 | (0.71-1.19) | 0.51031 | 1 |
| M46_other_inflammatory_spondylopathies | 97 | 5359 | 1.19 | (0.7-2.01) | 0.51594 | 1 |
| I37_pulmonary_valve_disorders | 36 | 5420 | 0.73 | (0.28-1.9) | 0.51796 | 1 |
| F40_phobic_anxiety_disorders | 81 | 5375 | 1.22 | (0.67-2.21) | 0.51887 | 1 |
| K26_duodenal_ulcer | 226 | 5230 | 0.89 | (0.61-1.28) | 0.52011 | 1 |
| F43_reaction_to_severe_stress_and_adjustment_disorders | 155 | 5301 | 0.85 | (0.53-1.39) | 0.52263 | 1 |
| M51_other_intervertebral_disk_disorders | 428 | 5028 | 0.91 | (0.69-1.21) | 0.52310 | 1 |
| F42_obsessive_compulsive_disorder | 12 | 5444 | 0.51 | (0.07-4.01) | 0.52311 | 1 |
| H72_perforation_of_tympanic_membrane | 34 | 5422 | 1.31 | (0.56-3.05) | 0.53255 | 1 |
| N12_tubulo_interstitial_nephritis_not_specified_as_acute_or_chronic | 68 | 5388 | 0.8 | (0.39-1.63) | 0.53374 | 1 |
| E03_other_hypothyroidism | 600 | 4856 | 1.08 | (0.85-1.37) | 0.53424 | 1 |
| N84_polyp_of_female_genital_tract | 153 | 5303 | 1.16 | (0.73-1.84) | 0.53953 | 1 |
| N42_other_disorders_of_prostate | 89 | 5367 | 1.17 | (0.7-1.97) | 0.54149 | 1 |
| J47_bronchiectasis | 250 | 5206 | 1.11 | (0.79-1.56) | 0.54528 | 1 |
| I74_arterial_embolism_and_thrombosis | 89 | 5367 | 1.18 | (0.69-2.03) | 0.54542 | 1 |
| N23_unspecified_renal_colic | 74 | 5382 | 1.2 | (0.66-2.17) | 0.55023 | 1 |
| G54_nerve_root_and_plexus_disorders | 29 | 5427 | 1.32 | (0.53-3.31) | 0.55146 | 1 |
| N45_orchitis_and_epididymitis | 54 | 5402 | 1.22 | (0.63-2.35) | 0.55163 | 1 |
| L20_atopic_dermatitis | 168 | 5288 | 1.13 | (0.75-1.7) | 0.55176 | 1 |
| G60_hereditary_and_idiopathic_neuropathy | 9 | 5447 | 1.65 | (0.32-8.61) | 0.55330 | 1 |
| J61_pneumoconiosis_due_to_asbestos_and_other_mineral_fibres | 44 | 5412 | 1.24 | (0.6-2.56) | 0.55580 | 1 |
| M12_other_specific_arthropathies | 19 | 5437 | 0.64 | (0.15-2.82) | 0.55635 | 1 |
| J81_pulmonary_oedema | 67 | 5389 | 1.2 | (0.65-2.19) | 0.56138 | 1 |
| L24_irritant_contact_dermatitis | 93 | 5363 | 0.84 | (0.48-1.49) | 0.56194 | 1 |
| E05_thyrotoxicosis_hyperthyroidism | 139 | 5317 | 0.86 | (0.52-1.43) | 0.56266 | 1 |
| H21_other_disorders_of_iris_and_ciliary_body | 30 | 5426 | 0.73 | (0.25-2.13) | 0.56391 | 1 |
| L92_granulomatous_disorders_of_skin_and_subcutaneous_tissue | 36 | 5420 | 1.28 | (0.55-2.98) | 0.56548 | 1 |
| E23_hypofunction_and_other_disorders_of_pituitary_gland | 38 | 5418 | 1.28 | (0.55-2.98) | 0.56720 | 1 |
| M45_ankylosing_spondylitis | 38 | 5418 | 0.76 | (0.29-1.97) | 0.57090 | 1 |
| E13_other_specified_diabetes_mellitus | 33 | 5423 | 1.29 | (0.53-3.16) | 0.57794 | 1 |
| G58_other_mononeuropathies | 17 | 5439 | 0.66 | (0.15-2.91) | 0.57931 | 1 |
| J45_asthma | 973 | 4483 | 0.95 | (0.78-1.15) | 0.58507 | 1 |
| H91_other_hearing_loss | 464 | 4992 | 0.93 | (0.71-1.21) | 0.59070 | 1 |
| E10_insulin_dependent_diabetes_mellitus | 214 | 5242 | 1.1 | (0.77-1.57) | 0.59373 | 1 |
| I36_nonrheumatic_tricuspid_valve_disorders | 25 | 5431 | 1.31 | (0.48-3.56) | 0.59973 | 1 |
| D61_other_aplastic_anaemias | 136 | 5320 | 0.88 | (0.53-1.44) | 0.60553 | 1 |
| H44_disorders_of_globe | 37 | 5419 | 0.78 | (0.3-2.03) | 0.61197 | 1 |
| H00_hordeolum_and_chalazion | 153 | 5303 | 1.12 | (0.72-1.73) | 0.61409 | 1 |
| H80_otosclerosis | 16 | 5440 | 0.68 | (0.15-3.06) | 0.61489 | 1 |
| M40_kyphosis_and_lordosis | 45 | 5411 | 0.78 | (0.3-2.03) | 0.61633 | 1 |
| G56_mononeuropathies_of_upper_limb | 361 | 5095 | 1.08 | (0.81-1.44) | 0.62021 | 1 |
| J98_other_respiratory_disorders | 881 | 4575 | 0.95 | (0.78-1.16) | 0.62081 | 1 |
| I95_hypotension | 689 | 4767 | 0.95 | (0.76-1.18) | 0.62413 | 1 |
| J94_other_pleural_conditions | 52 | 5404 | 1.2 | (0.58-2.5) | 0.62584 | 1 |
| F29_unspecified_nonorganic_psychosis | 15 | 5441 | 1.37 | (0.38-4.88) | 0.63129 | 1 |
| K52_other_non_infective_gastro_enteritis_and_colitis | 603 | 4853 | 1.06 | (0.84-1.34) | 0.63150 | 1 |
| F25_schizoaffective_disorders | 11 | 5445 | 0.6 | (0.08-4.83) | 0.63401 | 1 |
| H15_disorders_of_sclera | 19 | 5437 | 1.32 | (0.42-4.1) | 0.63689 | 1 |
| K42_umbilical_hernia | 190 | 5266 | 0.91 | (0.62-1.35) | 0.63715 | 1 |
| F80_specific_developmental_disorders_of_speech_and_language | 17 | 5439 | 0.7 | (0.16-3.12) | 0.64275 | 1 |
| N81_female_genital_prolapse | 214 | 5242 | 1.1 | (0.74-1.64) | 0.64560 | 1 |
| H40_glaucoma | 332 | 5124 | 0.93 | (0.69-1.27) | 0.64920 | 1 |
| H74_other_disorders_of_middle_ear_and_mastoid | 18 | 5438 | 0.71 | (0.16-3.17) | 0.65149 | 1 |
| N17_acute_renal_failure | 975 | 4481 | 0.96 | (0.79-1.16) | 0.66453 | 1 |
| N40_hyperplasia_of_prostate | 854 | 4602 | 1.05 | (0.85-1.29) | 0.66783 | 1 |
| M07_psoriatic_and_enteropathic_arthropathies | 29 | 5427 | 0.79 | (0.27-2.32) | 0.66852 | 1 |
| K27_peptic_ulcer_site_unspecified | 46 | 5410 | 1.18 | (0.56-2.48) | 0.67136 | 1 |
| N32_other_disorders_of_bladder | 506 | 4950 | 1.05 | (0.83-1.34) | 0.67303 | 1 |
| M53_other_dorsopathies_not_elsewhere_classified | 70 | 5386 | 1.15 | (0.61-2.17) | 0.67443 | 1 |
| K62_other_diseases_of_anus_and_rectum | 719 | 4737 | 1.05 | (0.85-1.3) | 0.67497 | 1 |
| F22_persistent_delusional_disorders | 26 | 5430 | 1.23 | (0.46-3.33) | 0.67920 | 1 |
| J43_emphysema | 343 | 5113 | 1.06 | (0.79-1.44) | 0.68483 | 1 |
| G61_inflammatory_polyneuropathy | 24 | 5432 | 0.77 | (0.23-2.67) | 0.68555 | 1 |
| H73_other_disorders_of_tympanic_membrane | 14 | 5442 | 0.73 | (0.16-3.32) | 0.68694 | 1 |
| F45_somatoform_disorders | 29 | 5427 | 0.81 | (0.28-2.36) | 0.69457 | 1 |
| D51_vitamin_b12_deficiency_anaemia | 102 | 5354 | 0.9 | (0.53-1.54) | 0.69810 | 1 |
| J86_pyothorax | 25 | 5431 | 1.21 | (0.45-3.28) | 0.70327 | 1 |
| N05_unspecified_nephritic_syndrome | 29 | 5427 | 0.82 | (0.28-2.37) | 0.70792 | 1 |
| N03_chronic_nephritic_syndrome | 43 | 5413 | 1.15 | (0.54-2.45) | 0.71204 | 1 |
| K72_hepatic_failure_not_elsewhere_classified | 108 | 5348 | 1.1 | (0.66-1.82) | 0.71233 | 1 |
| G80_infantile_cerebral_palsy | 5 | 5451 | 1.52 | (0.15-15.35) | 0.72048 | 1 |
| H46_optic_neuritis | 13 | 5443 | 0.69 | (0.09-5.38) | 0.72162 | 1 |
| I07_rheumatic_tricuspid_valve_diseases | 85 | 5371 | 1.11 | (0.62-1.96) | 0.72806 | 1 |
| H53_visual_disturbances | 292 | 5164 | 0.94 | (0.68-1.31) | 0.73384 | 1 |
| K85_acute_pancreatitis | 127 | 5329 | 1.08 | (0.67-1.74) | 0.74047 | 1 |
| I69_sequelae_of_cerebrovascular_disease | 208 | 5248 | 1.07 | (0.73-1.55) | 0.74162 | 1 |
| F50_eating_disorders | 9 | 5447 | 1.41 | (0.17-11.6) | 0.74696 | 1 |
| I08_multiple_valve_diseases | 356 | 5100 | 1.05 | (0.79-1.39) | 0.74714 | 1 |
| J46_status_asthmaticus | 10 | 5446 | 1.29 | (0.27-6.19) | 0.74800 | 1 |
| L28_lichen_simplex_chronicus_and_prurigo | 21 | 5435 | 0.82 | (0.24-2.83) | 0.74803 | 1 |
| N75_diseases_of_bartholin's_gland | 15 | 5441 | 1.28 | (0.28-5.88) | 0.74805 | 1 |
| N30_cystitis | 268 | 5188 | 1.06 | (0.75-1.49) | 0.75667 | 1 |
| H27_other_disorders_of_lens | 18 | 5438 | 1.22 | (0.35-4.28) | 0.75738 | 1 |
| J92_pleural_plaque | 142 | 5314 | 1.07 | (0.7-1.64) | 0.76634 | 1 |
| I27_other_pulmonary_heart_diseases | 198 | 5258 | 0.94 | (0.64-1.39) | 0.76795 | 1 |
| I72_other_aneurysm | 54 | 5402 | 0.9 | (0.43-1.86) | 0.76828 | 1 |
| N62_hypertrophy_of_breast | 36 | 5420 | 0.87 | (0.33-2.27) | 0.76869 | 1 |
| N64_other_disorders_of_breast | 121 | 5335 | 0.92 | (0.53-1.61) | 0.77601 | 1 |
| H55_nystagmus_and_other_irregular_eye_movements | 18 | 5438 | 1.19 | (0.33-4.23) | 0.79054 | 1 |
| H11_other_disorders_of_conjunctiva | 115 | 5341 | 0.93 | (0.56-1.57) | 0.79717 | 1 |
| G83_other_paralytic_syndromes | 53 | 5403 | 1.1 | (0.54-2.21) | 0.79805 | 1 |
| K74_fibrosis_and_cirrhosis_of_liver | 170 | 5286 | 0.95 | (0.62-1.44) | 0.79857 | 1 |
| K80_cholelithiasis | 617 | 4839 | 0.97 | (0.77-1.22) | 0.79891 | 1 |
| I48_atrial_fibrillation_and_flutter | 1450 | 4006 | 0.98 | (0.83-1.16) | 0.80032 | 1 |
| L30_other_dermatitis | 643 | 4813 | 1.03 | (0.82-1.29) | 0.80598 | 1 |
| G45_transient_cerebral_ischaemic_attacks_and_related_syndromes | 296 | 5160 | 1.04 | (0.76-1.43) | 0.80860 | 1 |
| N47_redundant_prepuce_phimosis_and_paraphimosis | 102 | 5354 | 0.94 | (0.56-1.58) | 0.82083 | 1 |
| K46_unspecified_abdominal_hernia | 57 | 5399 | 1.08 | (0.54-2.18) | 0.82316 | 1 |
| L21_seborrhoeic_dermatitis | 137 | 5319 | 1.05 | (0.66-1.69) | 0.82466 | 1 |
| N18_chronic_renal_failure | 1112 | 4344 | 1.02 | (0.85-1.22) | 0.83032 | 1 |
| N94_pain_and_other_conditions_associated_with_female_genital_organs_and_menstrual_cycle | 78 | 5378 | 1.08 | (0.53-2.21) | 0.83120 | 1 |
| I60_subarachnoid_haemorrhage | 37 | 5419 | 0.9 | (0.35-2.35) | 0.83357 | 1 |
| G24_dystonia | 26 | 5430 | 1.12 | (0.37-3.37) | 0.83562 | 1 |
| M18_arthrosis_of_first_carpometacarpal_joint | 33 | 5423 | 0.9 | (0.34-2.41) | 0.83921 | 1 |
| I47_paroxysmal_tachycardia | 170 | 5286 | 0.96 | (0.63-1.46) | 0.84293 | 1 |
| G43_migraine | 251 | 5205 | 1.04 | (0.72-1.5) | 0.85167 | 1 |
| E04_other_non_toxic_goitre | 85 | 5371 | 0.94 | (0.51-1.73) | 0.85168 | 1 |
| I12_hypertensive_renal_disease | 119 | 5337 | 1.05 | (0.65-1.68) | 0.85388 | 1 |
| E29_testicular_dysfunction | 14 | 5442 | 0.87 | (0.19-3.92) | 0.85669 | 1 |
| G35_multiple_sclerosis | 58 | 5398 | 1.08 | (0.48-2.44) | 0.85931 | 1 |
| K44_diaphragmatic_hernia | 1028 | 4428 | 0.98 | (0.82-1.19) | 0.86518 | 1 |
| F60_specific_personality_disorders | 20 | 5436 | 0.9 | (0.26-3.13) | 0.86805 | 1 |
| H26_other_cataract | 980 | 4476 | 1.02 | (0.84-1.23) | 0.87261 | 1 |
| I63_cerebral_infarction | 311 | 5145 | 0.98 | (0.71-1.33) | 0.87693 | 1 |
| N60_benign_mammary_dysplasia | 104 | 5352 | 0.95 | (0.52-1.74) | 0.87822 | 1 |
| M31_other_necrotising_vasculopathies | 49 | 5407 | 1.06 | (0.49-2.3) | 0.88231 | 1 |
| L25_unspecified_contact_dermatitis | 35 | 5421 | 0.93 | (0.36-2.44) | 0.88512 | 1 |
| G40_epilepsy | 254 | 5202 | 1.03 | (0.72-1.46) | 0.88918 | 1 |
| I51_complications_and_ill_defined_descriptions_of_heart_disease | 546 | 4910 | 0.98 | (0.78-1.25) | 0.89076 | 1 |
| M32_systemic_lupus_erythematosus | 29 | 5427 | 0.93 | (0.32-2.73) | 0.89178 | 1 |
| N34_urethritis_and_urethral_syndrome | 17 | 5439 | 1.09 | (0.3-3.95) | 0.89570 | 1 |
| I71_aortic_aneurysm_and_dissection | 182 | 5274 | 1.03 | (0.69-1.53) | 0.89632 | 1 |
| L89_decubitus_ulcer | 558 | 4898 | 1.02 | (0.8-1.29) | 0.90309 | 1 |
| N73_other_female_pelvic_inflammatory_diseases | 56 | 5400 | 1.05 | (0.48-2.28) | 0.90345 | 1 |
| N36_other_disorders_of_urethra | 35 | 5421 | 0.94 | (0.36-2.5) | 0.90912 | 1 |
| L23_allergic_contact_dermatitis | 10 | 5446 | 1.09 | (0.23-5.26) | 0.91427 | 1 |
| N20_calculus_of_kidney_and_ureter | 194 | 5262 | 1.02 | (0.69-1.5) | 0.91518 | 1 |
| I42_cardiomyopathy | 106 | 5350 | 0.97 | (0.58-1.63) | 0.92279 | 1 |
| N61_inflammatory_disorders_of_breast | 17 | 5439 | 0.93 | (0.21-4.17) | 0.92501 | 1 |
| K76_other_diseases_of_liver | 507 | 4949 | 0.99 | (0.77-1.27) | 0.93198 | 1 |
| K22_other_diseases_of_oesophagus | 499 | 4957 | 1.01 | (0.78-1.3) | 0.93479 | 1 |
| K60_fissure_and_fistula_of_anal_and_rectal_regions | 89 | 5367 | 0.98 | (0.55-1.73) | 0.93500 | 1 |
| F10_mental_and_behavioural_disorders_due_to_use_of_alcohol | 416 | 5040 | 0.99 | (0.75-1.31) | 0.93960 | 1 |
| M48_other_spondylopathies | 408 | 5048 | 1.01 | (0.77-1.33) | 0.93995 | 1 |
| M16_coxarthrosis_arthrosis_of_hip | 480 | 4976 | 1.01 | (0.78-1.31) | 0.94349 | 1 |
| E28_ovarian_dysfunction | 16 | 5440 | 0.95 | (0.21-4.27) | 0.94469 | 1 |
| H57_other_disorders_of_eye_and_adnexa | 114 | 5342 | 1.02 | (0.61-1.69) | 0.94640 | 1 |
| L50_urticaria | 113 | 5343 | 1.02 | (0.6-1.73) | 0.94790 | 1 |
| I24_other_acute_ischaemic_heart_diseases | 147 | 5309 | 1.01 | (0.66-1.56) | 0.94804 | 1 |
| K21_gastro_oesophageal_reflux_disease | 1272 | 4184 | 0.99 | (0.84-1.18) | 0.94919 | 1 |
| I45_other_conduction_disorders | 337 | 5119 | 0.99 | (0.74-1.33) | 0.95381 | 1 |
| G62_other_polyneuropathies | 250 | 5206 | 1.01 | (0.72-1.42) | 0.95603 | 1 |
| G91_hydrocephalus | 76 | 5380 | 1.02 | (0.53-1.96) | 0.96114 | 1 |
| N72_inflammatory_disease_of_cervix_uteri | 19 | 5437 | 1.03 | (0.29-3.62) | 0.96861 | 1 |
| J96_respiratory_failure_not_elsewhere_classified | 323 | 5133 | 0.99 | (0.73-1.36) | 0.96933 | 1 |
| K41_femoral_hernia | 22 | 5434 | 1.02 | (0.3-3.48) | 0.97677 | 1 |
| K63_other_diseases_of_intestine | 725 | 4731 | 1 | (0.8-1.24) | 0.97995 | 1 |
| G57_mononeuropathies_of_lower_limb | 62 | 5394 | 0.99 | (0.5-1.98) | 0.98024 | 1 |
| N76_other_inflammation_of_vagina_and_vulva | 46 | 5410 | 1.01 | (0.42-2.43) | 0.98844 | 1 |
| M13_other_arthritis | 811 | 4645 | 1 | (0.82-1.23) | 0.99082 | 1 |
| K90_intestinal_malabsorption | 67 | 5389 | 1 | (0.5-1.98) | 0.99125 | 1 |
